# Systemic inflammatory syndromes as life-threatening side effects of immune checkpoint inhibitors: Systematic review of the literature

**DOI:** 10.1101/2022.07.28.22278028

**Authors:** Lisa L. Liu, Marcus Skribek, Ulrika Harmenberg, Marco Gerling

## Abstract

Immune checkpoint inhibitors (ICIs) are associated with a wide range of immune-related adverse events (irAEs). As oncological indications for ICIs widen, their rare side effects become increasingly visible in clinical practice and impact therapy decisions.

Here, we provide a systematic review of the literature of CRS and related life-threatening side effects of ICI treatment, such as hemophagocytic lymphohistiocytosis (HLH). We searched Medline, Embase, and the Web of Science Core Collection from inception until October 2021 for reports on CRS, cytokine storm, macrophage activation syndrome, HLH, and related hyperinflammatory disorders in patients with solid cancers receiving ICIs. We found *n* = 1866 articles, which were assessed for eligibility independently by two examiners. Of those, *n* = 49 articles reporting on *n* = 189 individuals were eligible for review. We found that the median time from last infusion to the occurrence of CRS/HLH was approximately nine days, while the onset of symptoms varied from immediately after infusion to one month after treatment. Most patients were treated with either corticosteroids or the anti-interleukin 6 (IL-6) antibody tocilizumab, and although the majority of patients recovered, a few cases were fatal. Concomitant IL-6 and ICI treatment was reported as beneficial for both the antitumoral effect and for limiting side effects. Data from international pharmacovigilance databases underscored that ICI-related CRS and HLH are rare events, but we identified significant differences in reported frequencies, which might suggest substantial underreporting.

The results from this first systematic review of CRS/HLH due to ICI therapy highlight that life-threatening systemic inflammatory complications of ICIs are rare and non-fatal in the majority of patients. Limited data support the use of IL-6 inhibitors in combination with ICIs to augment the antitumoral effect and reduce hyperinflammation.

## Introduction

Immune checkpoint inhibitors (ICIs) have become important therapeutic options for various tumor types. ICIs are associated with specific toxicities, commonly referred to as immune-related adverse events (irAEs), which can lead to treatment interruption or discontinuation. International guidelines aid clinicians in the diagnosis and management of relatively common irAEs, such as skin rashes, colitis, thyroiditis, and pneumonitis^1,2^. However, the increasing volume of patients treated with ICIs is starting to reveal less common side effects, including systemic hyperinflammatory syndromes. Nonspecific systemic inflammatory reactions to ICIs, such as self-limiting fever or skin rashes during or shortly after infusion^2^, need to be distinguished from severe, persistent, and potentially life-threatening conditions such as cytokine release syndrome (CRS) and hemophagocytic lymphohistiocytosis (HLH)/macrophage-activation syndrome (MAS)^3^. Due to their severity, these irAEs are of particular clinical importance and require a decision on continuation of ICI treatment, for which evidence is lacking. Because of their rarity, hyperinflammatory syndrome are incompletely captured in randomized clinical trials with ICIs ^4,5^, and not yet discussed in irAE guidelines^1,2^. Hence, real-world data and case reports of rare irAEs are needed to understand their frequency and severity, and to improve clinical management.

In cancer therapy, CRS is best understood in the context of chimeric antigen receptor (CAR) T cell therapies, where it occurs in a substantial proportion of patients at different levels of severity^6^. CRS is believed to be mainly driven by T cell-derived interferon gamma (IFN-γ), which stimulates macrophages to produce various pro-inflammatory substances including interleukin 6 (IL-6) and tumor necrosis factor alpha (TNF-α)^7^. Therapeutically, IL-6 inhibition with specific anti-IL-6 receptor antibodies, such as tocilizumab, has proven highly effective against CRS, reflected by the US Federal Drug Agency’s approval of tocilizumab for CAR T cell-induced CRS^8^. This is important because IL-6 inhibition could allow for the continuation of any treatment associated with mild CRS^9^.

HLH is an umbrella term for life-threatening hyperinflammatory conditions with supramaximal activation of the immune system. For the diagnosis of HLH, the HLH-2004 diagnostic criteria are frequently used, although they were developed for the pediatric population^3^. The criteria include clinical features such as fever and splenomegaly, as well as laboratory findings such as cytopenias, hypertriglyceridemia, hyperferritinemia, evidence of hemophagocytosis, and the absence of natural killer (NK) cell activity^3^. More recently, the HScore, which includes similar criteria to HLH-2004, was developed to estimate the probability for reactive HLH in adults with inflammatory syndromes^10^. Genetic analyses from pediatric patients have revealed a wide variety of predisposing variants that presumably play a role for different immune cell types, suggesting that HLH-related conditions represent a complex disease continuum^11^.

Reports on hyperinflammatory syndromes due to ICI treatment have started to emerge during recent years, and have suggested that these are relatively rare, but potentially life-threatening events^12^. Both HLH and CRS can be fatal by causing hypotension, capillary leak syndrome, and consequently organ dysfunction^7^. Because of the potential risk of increasing the severity of CRS/HLH upon repeated exposure to a particular trigger, suspicion of these hyperinflammatory syndromes in clinical practice most often leads to treatment cessation. Hence, a better understanding of this complex disease spectrum in the context of ICI treatment is needed to guide decision-making on treatment continuation and optimal management.

To set the stage for a more evidence-based approach to CRS, we conducted a systematic review of the literature, in which we identify *n* = 49 articles on *n* = 189 patients with hyperinflammatory syndromes due to ICIs. The results reveal that most patients with CRS/HLH recover and that a fatal outcome is rare. In addition, the literature reveals that corticosteroids and IL-6 inhibition may provide effective therapies. Extrapolating from preclinical data, which we review in brief, we posit that rechallenging with ICIs after CRS/HLH has been controlled should be considered at least in patients with mild hyperinflammatory ICI side effects who are expected to benefit from ICI treatment. Recent data suggest that ICIs in combination with IL-6 antagonists may boost the antitumoral effect, while simultaneously protecting from severe irAEs, which makes ICIs in combination with IL-6 inhibition an attractive option for rechallenge that warrants further research.

## Methods

### Systematic review

We performed a systematic search for published reports on CRS, HLH and related hyperinflammatory diseases in MEDLINE, Embase, and the Web of Science databases as of October 2021. The search strategies were designed in collaboration with the Karolinska Institute library and are included as **Supplementary Figure 1**. Deduplication was performed as previously described^13^.

Additional manual searches were performed based on the lists of references in the eligible studies, and a reduced set of keywords in the MEDLINE database only, as of June 13^th^ 2022. Inclusion criteria for abstract review were assessed independently by two reviewers (LLL and MG) and defined as follows: (1) report on any hyperinflammatory syndrome (CRS, HLH, MAS, or any indication of these or related systemic syndromes) on human patients with solid tumors, (2) reported use of any ICI at any time of the treatment, (3) case series or case reports, i.e. no randomized-controlled trial (RCT), unless explicitly stated that CRS/HLH/MAS or any other hyperinflammatory syndrome was reported, (4) articles in English, Swedish, Chinese or German. LLL and MG agreed on article inclusion by discussing the articles for which the initial decision on inclusion differed. For all eligible articles, the full-text was downloaded by LLL or MG. Data were extracted by either LLL, MS, or MG, and all authors convened to agree on final inclusion. As the majority of the included studies were case reports on rare events, no further criteria for study quality assessment were applied and the risk for bias was not assessed. The systematic review was not preregistered.

## Results

### Systematic review

To chart our current knowledge of hyperinflammatory syndromes, such as CRS and HLH, as rare side effects of treatment with ICIs, we conducted a systematic review of the literature housed in the major medical databases as of October 2021. In addition, we manually updated the list of eligible studies as of 13^th^ June 2022. **Supplementary Figure 2** presents the PRISMA reporting chart for the study.

**Supplementary Figure 2.**
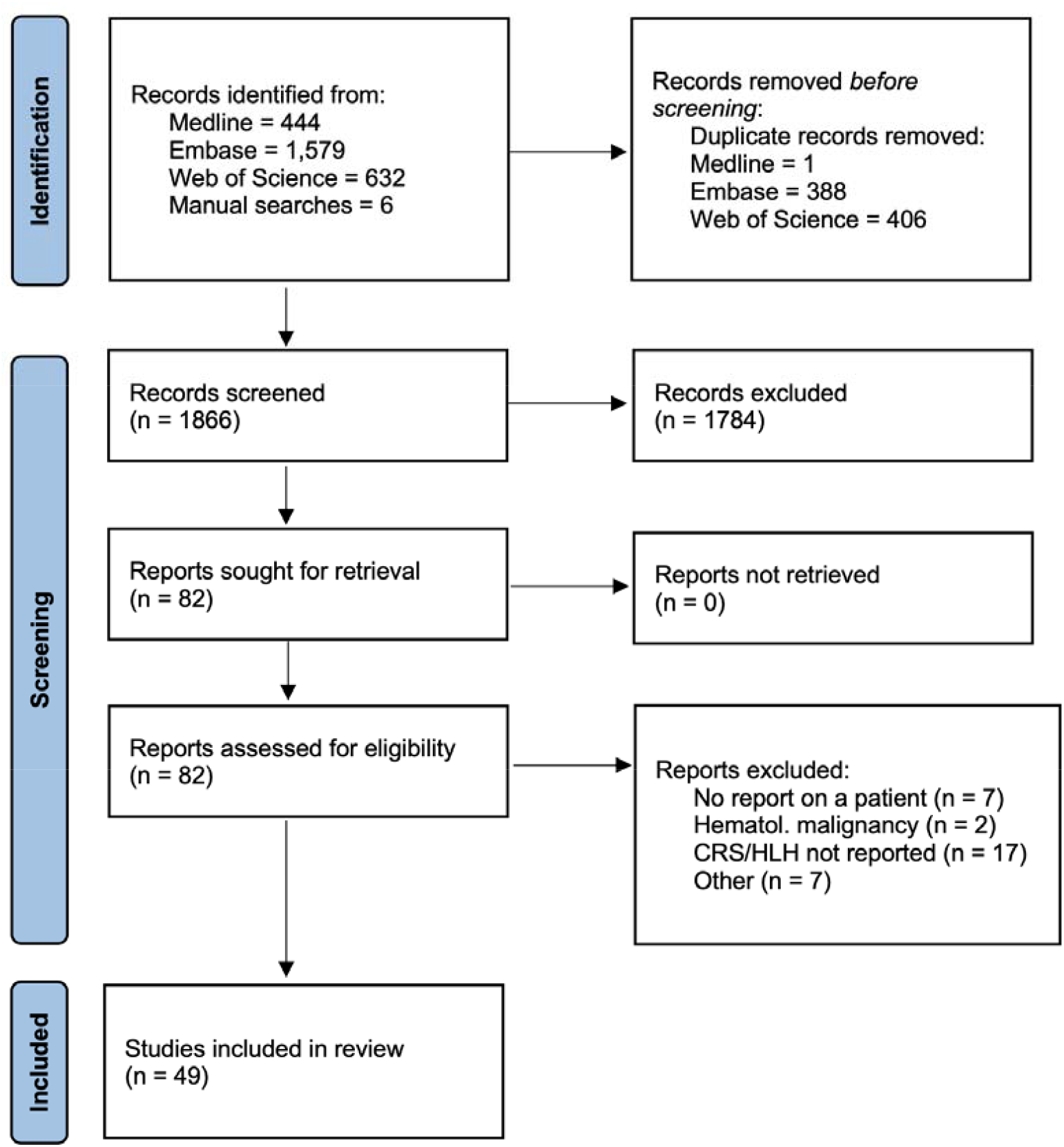
PRISMA 2020 flow diagram of the study.

We identified a total of *n* = 49 articles, reporting on *n* = 189 patients treated in the USA, the UK, Switzerland, Spain, Poland, Lebanon, Japan, Israel, Germany, France, China, Canada, Australia, and Singapore (**Table 1**). Of these studies, *n* = 45 reports were on *n* = < 5 individual patients (comprising a total of *n* = 56 individuals), while *n* = 4 studies reported case series or data from queries of pharmacovigilance databases (**Table 1**). Definitions of CRS/HLH varied significantly between reports, and retrospective diagnostic assessment was not possible due to partially incomplete and heterogeneous clinical data. Therefore, we used the respective authors’ assessment of the hyperinflammatory condition as CRS, HLH, or related hyperinflammatory diseases for further analysis. The earliest published report was from 2016^12^.

**Table 1.**
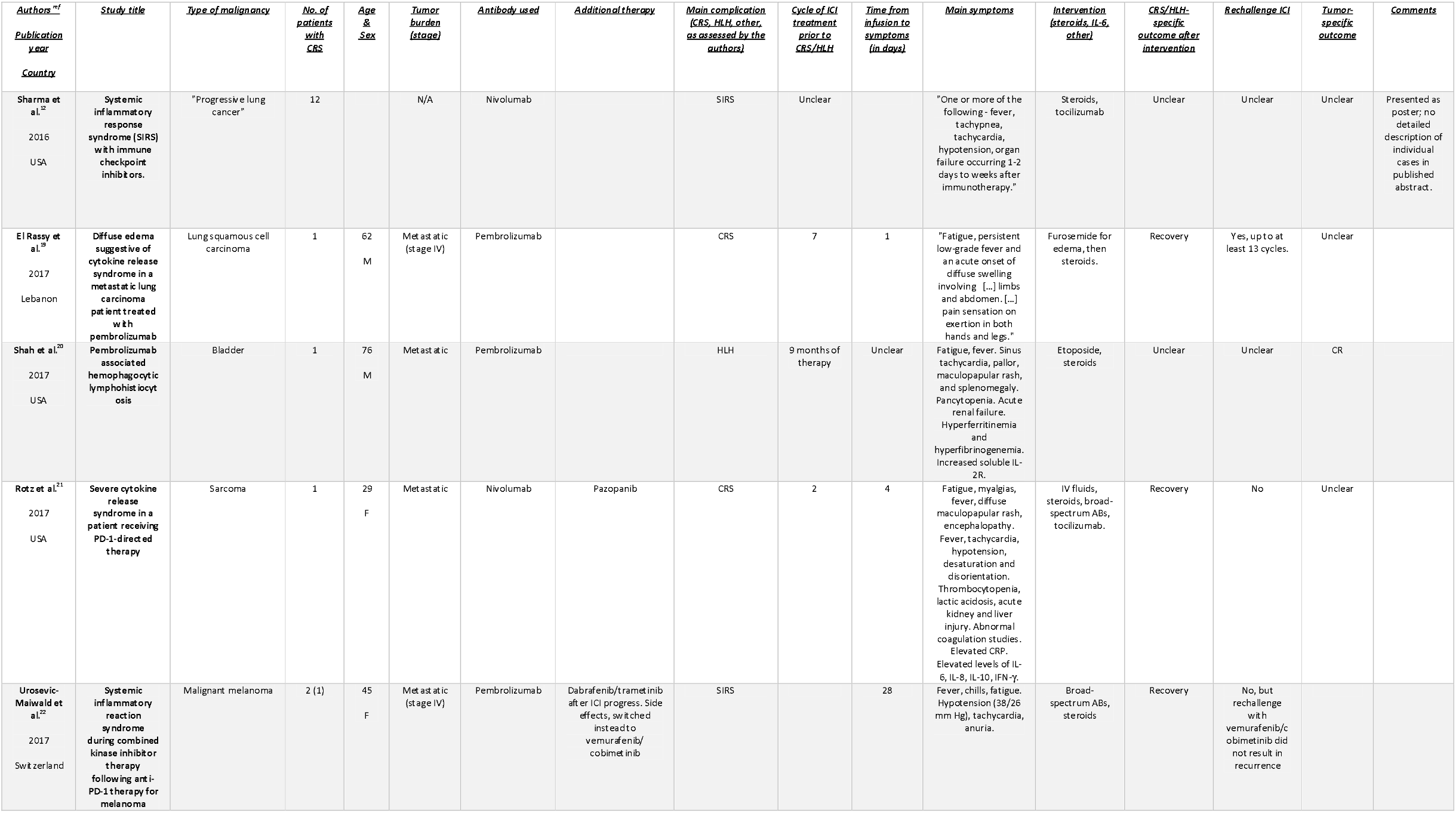
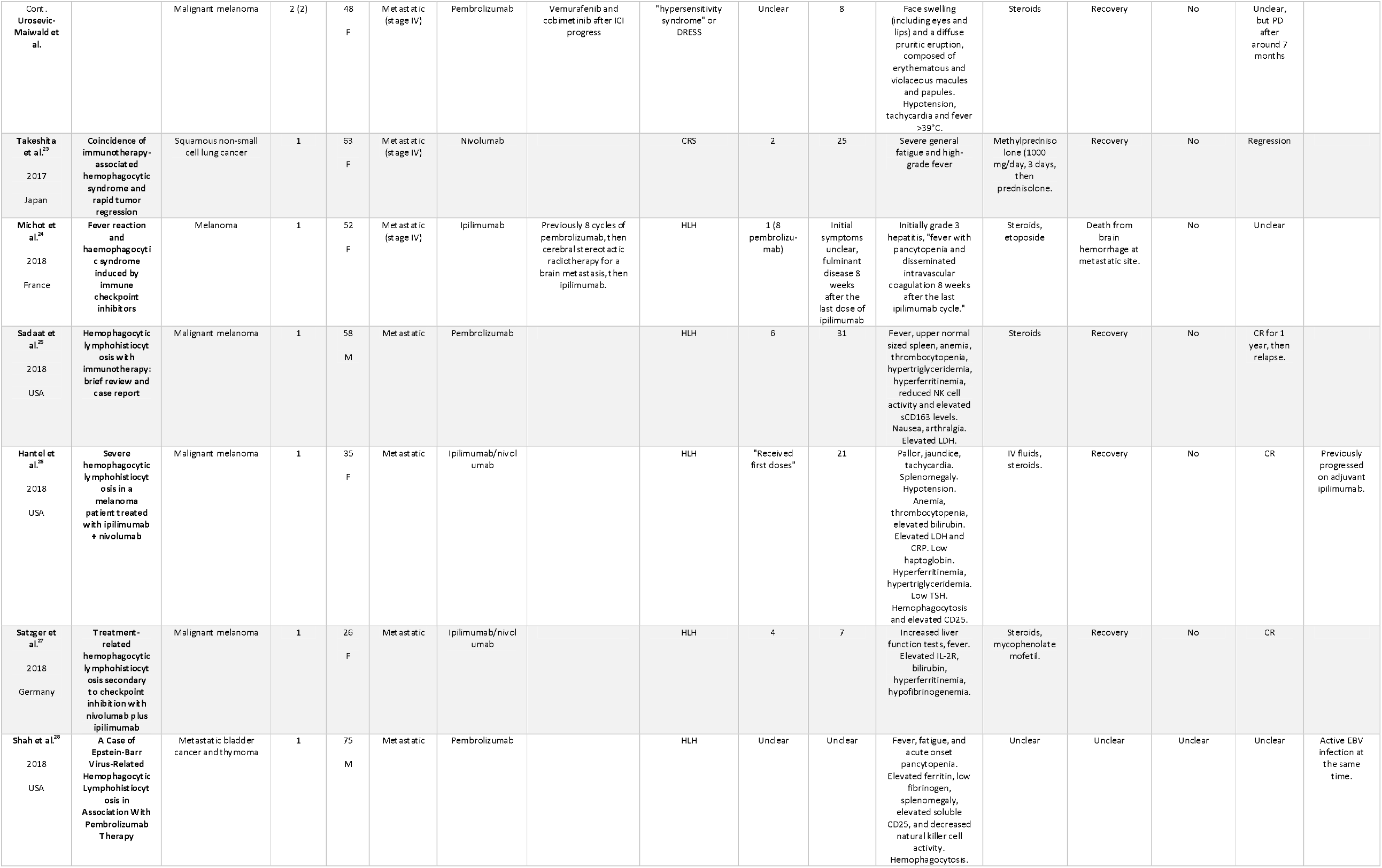
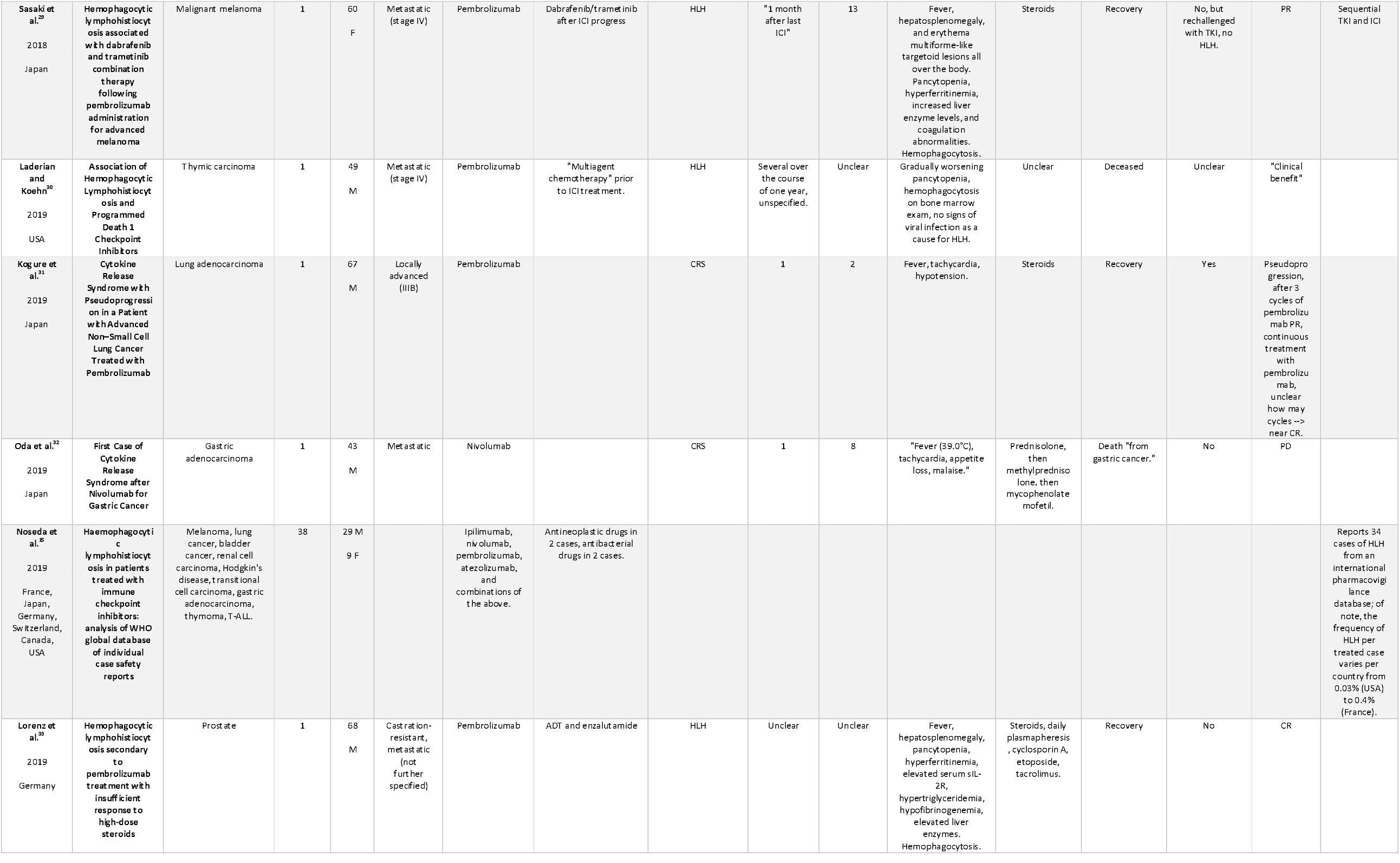
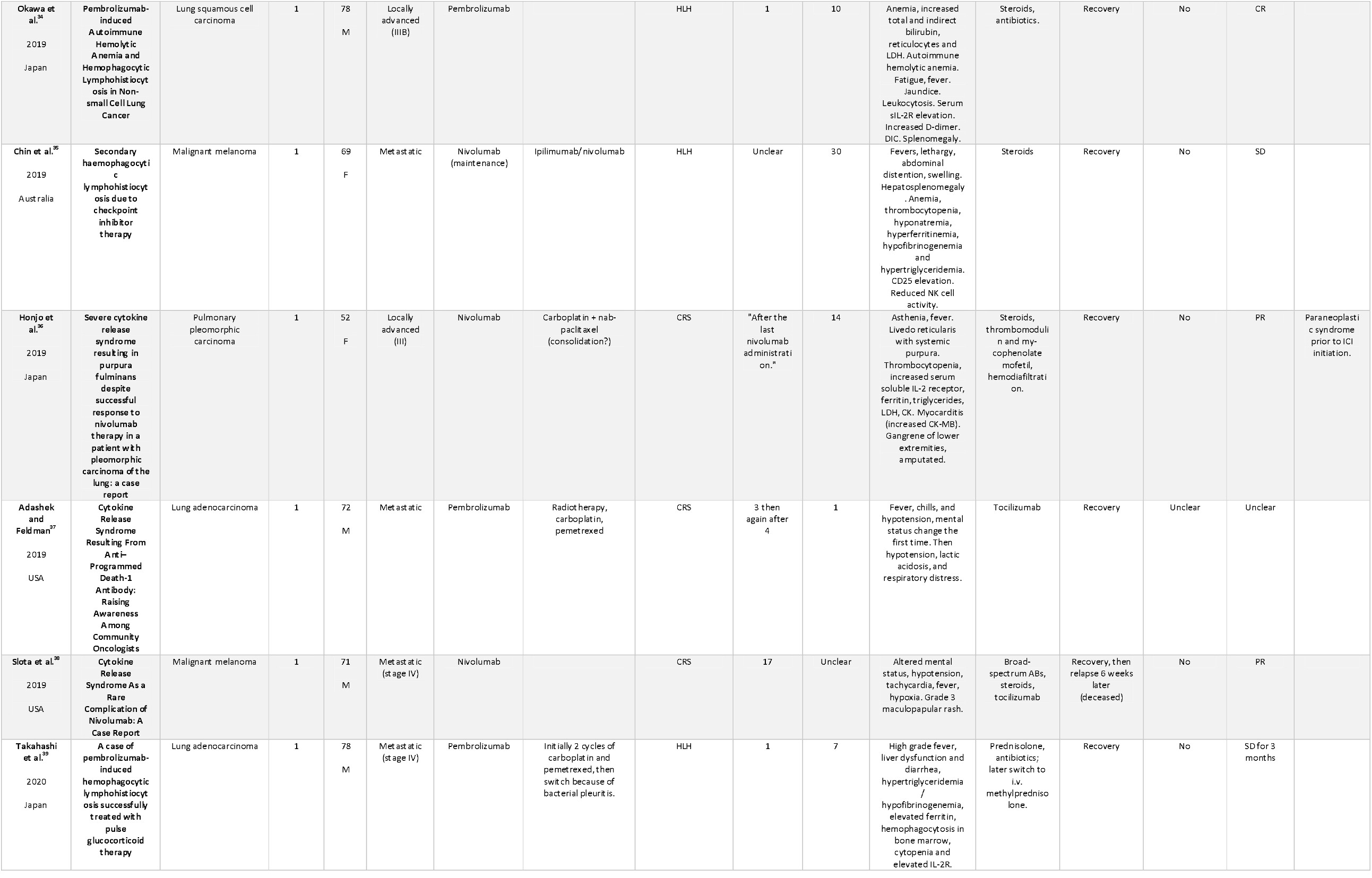
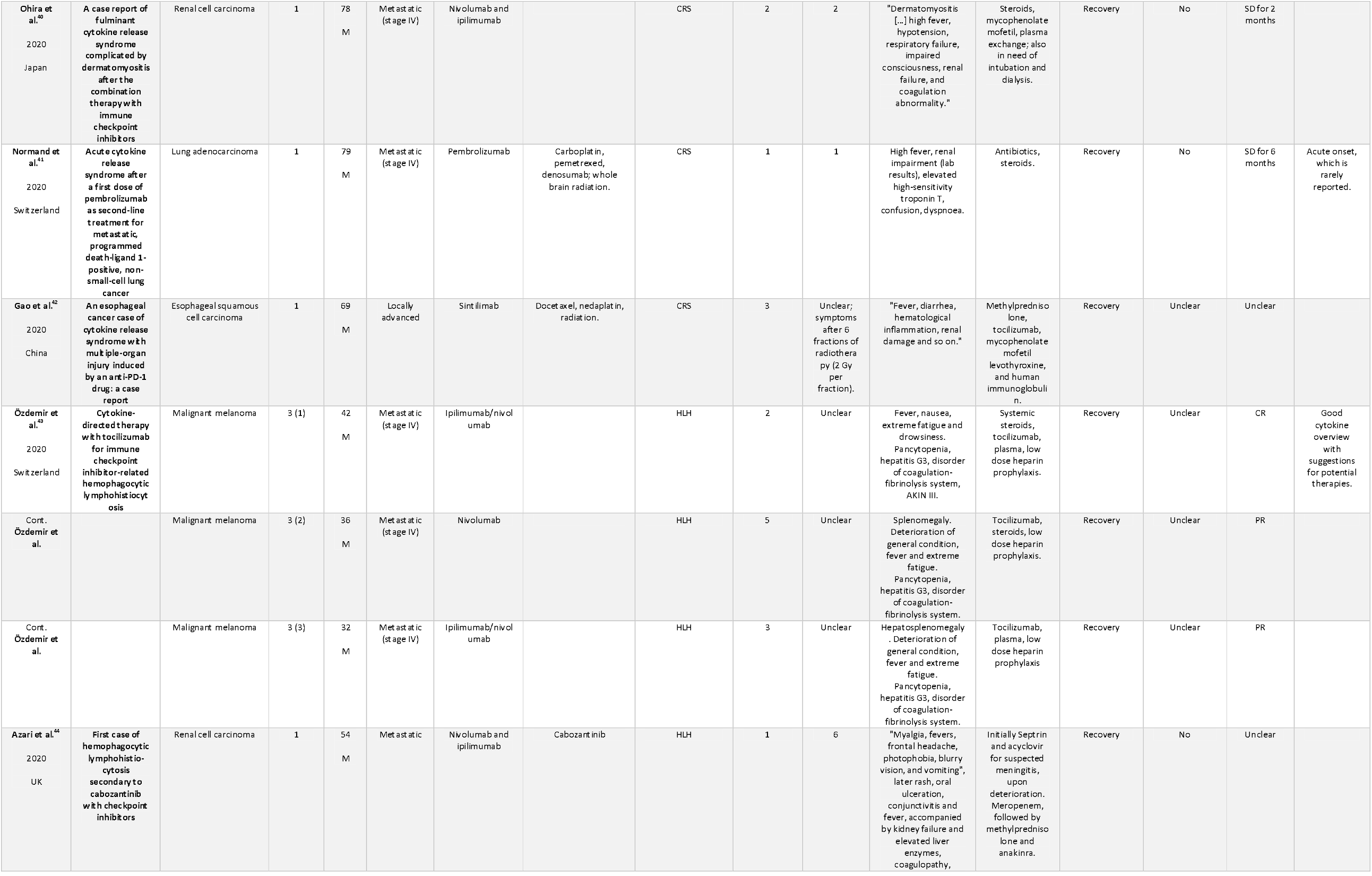
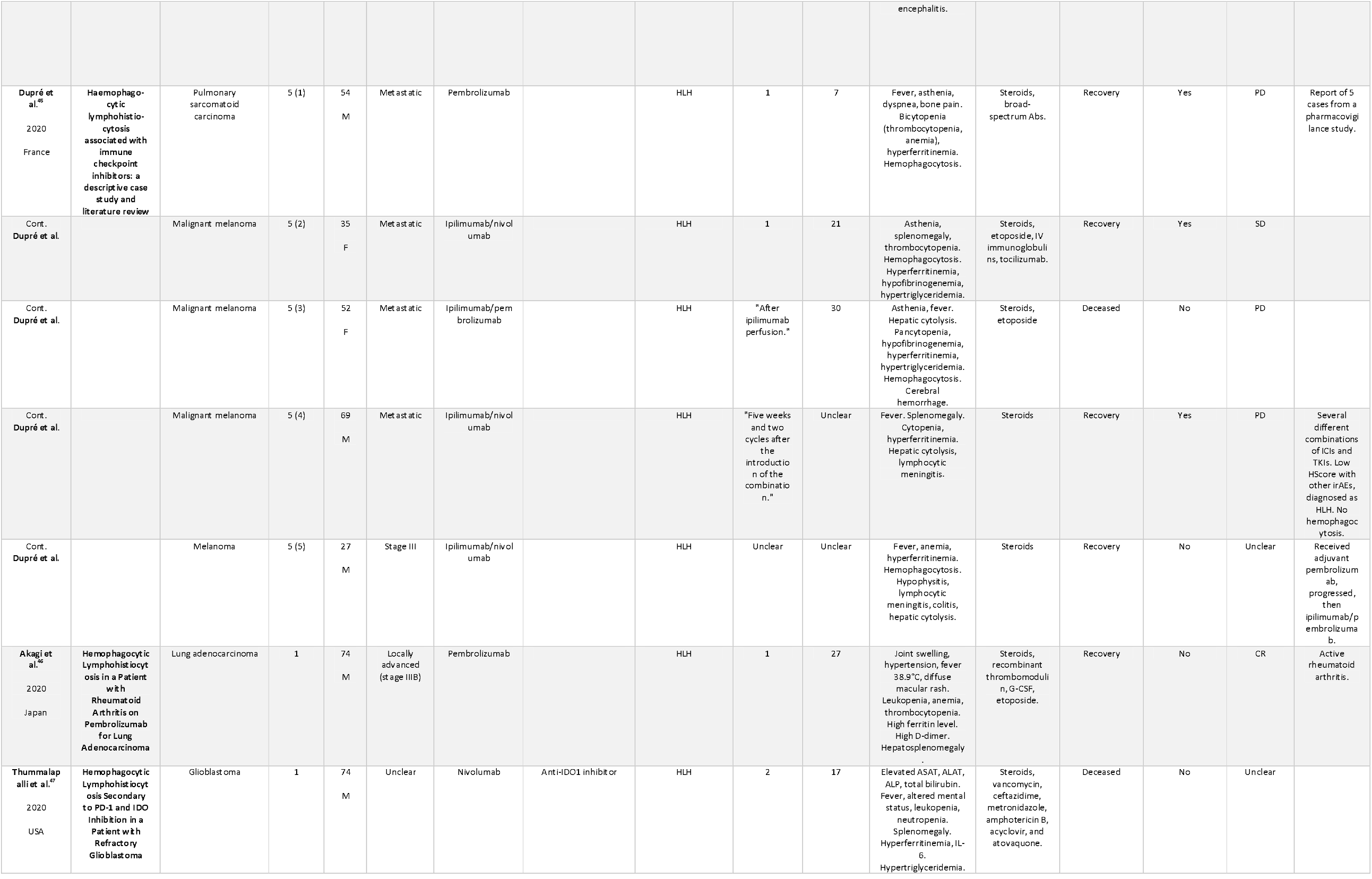
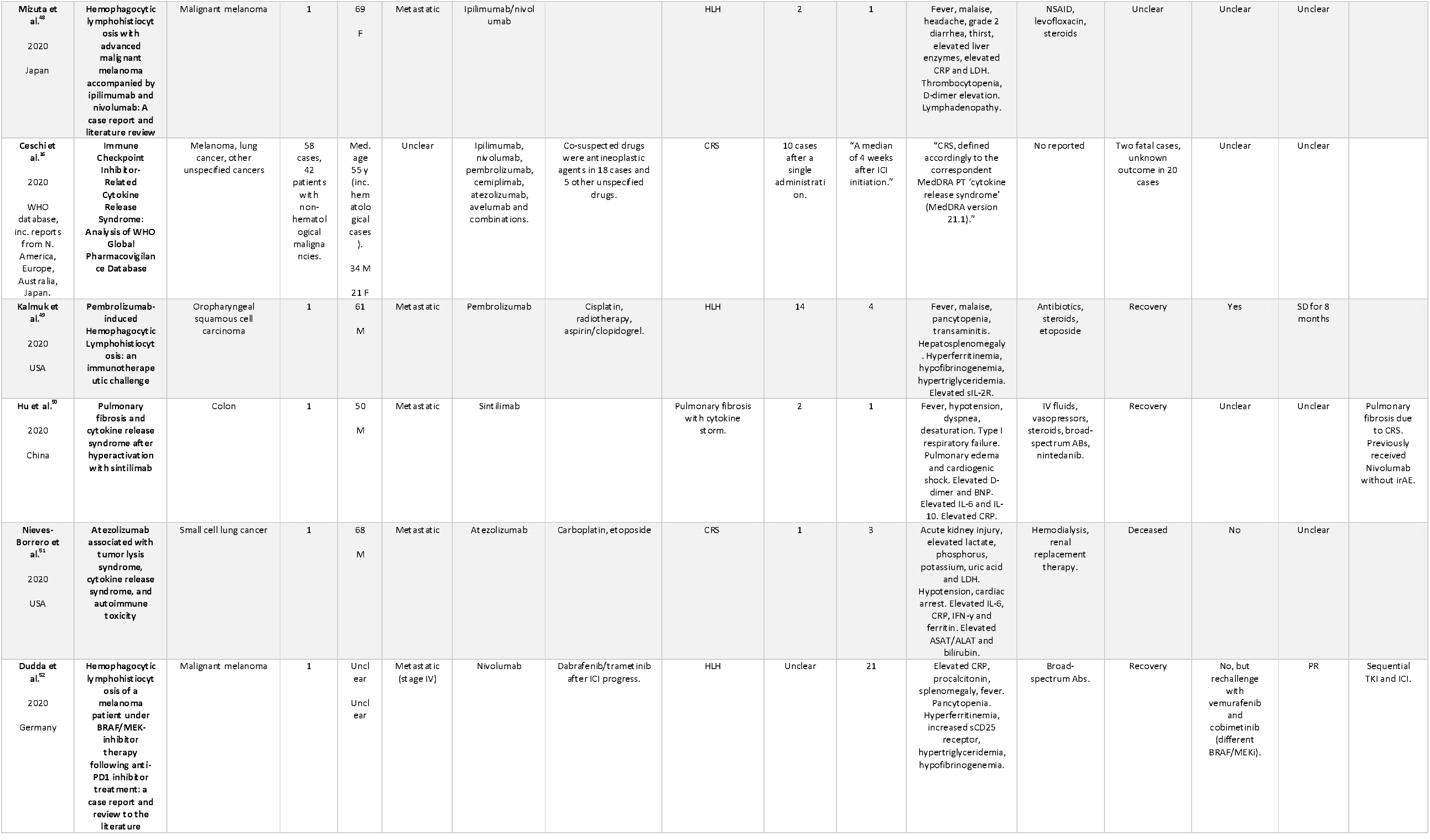
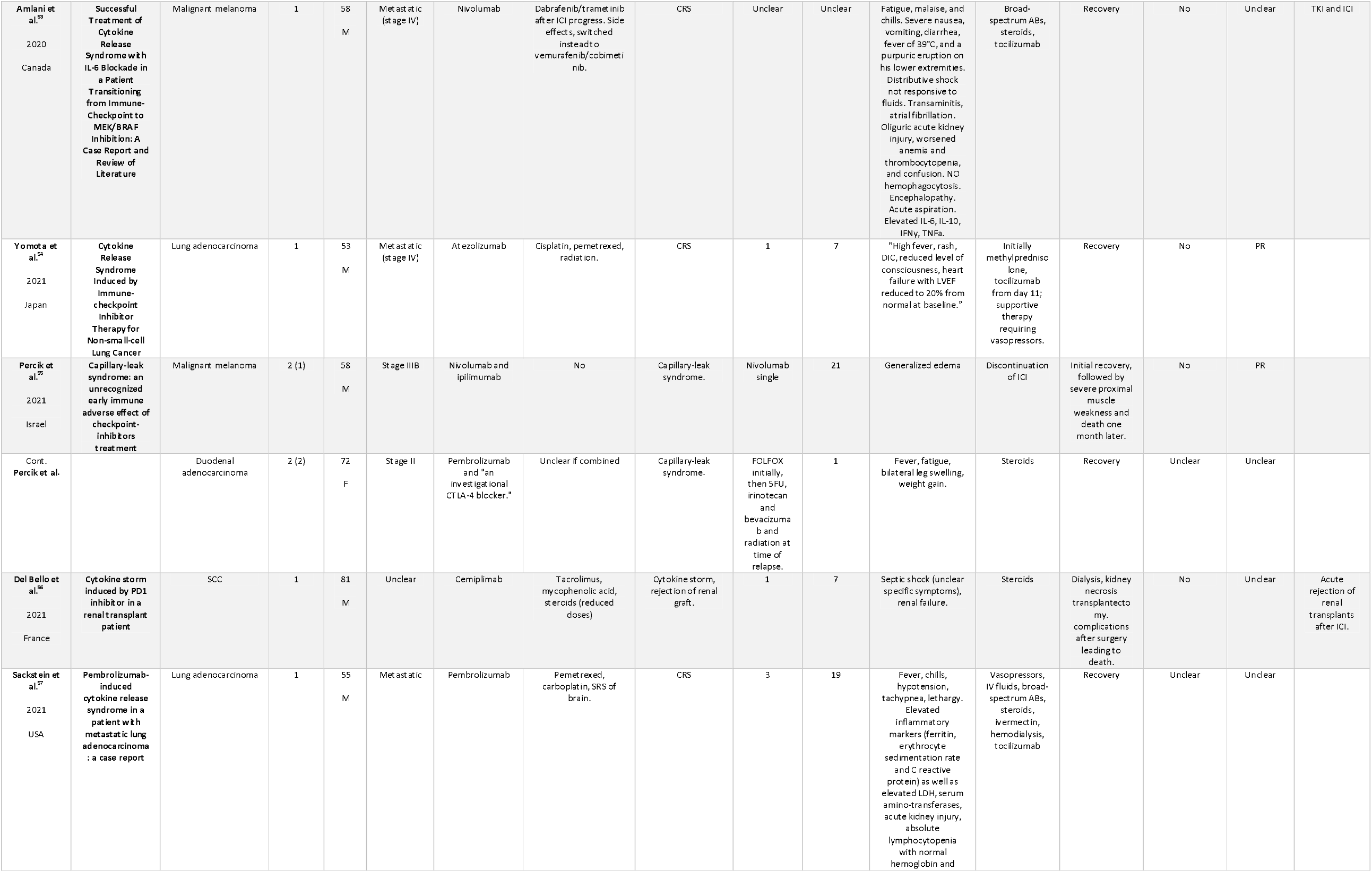
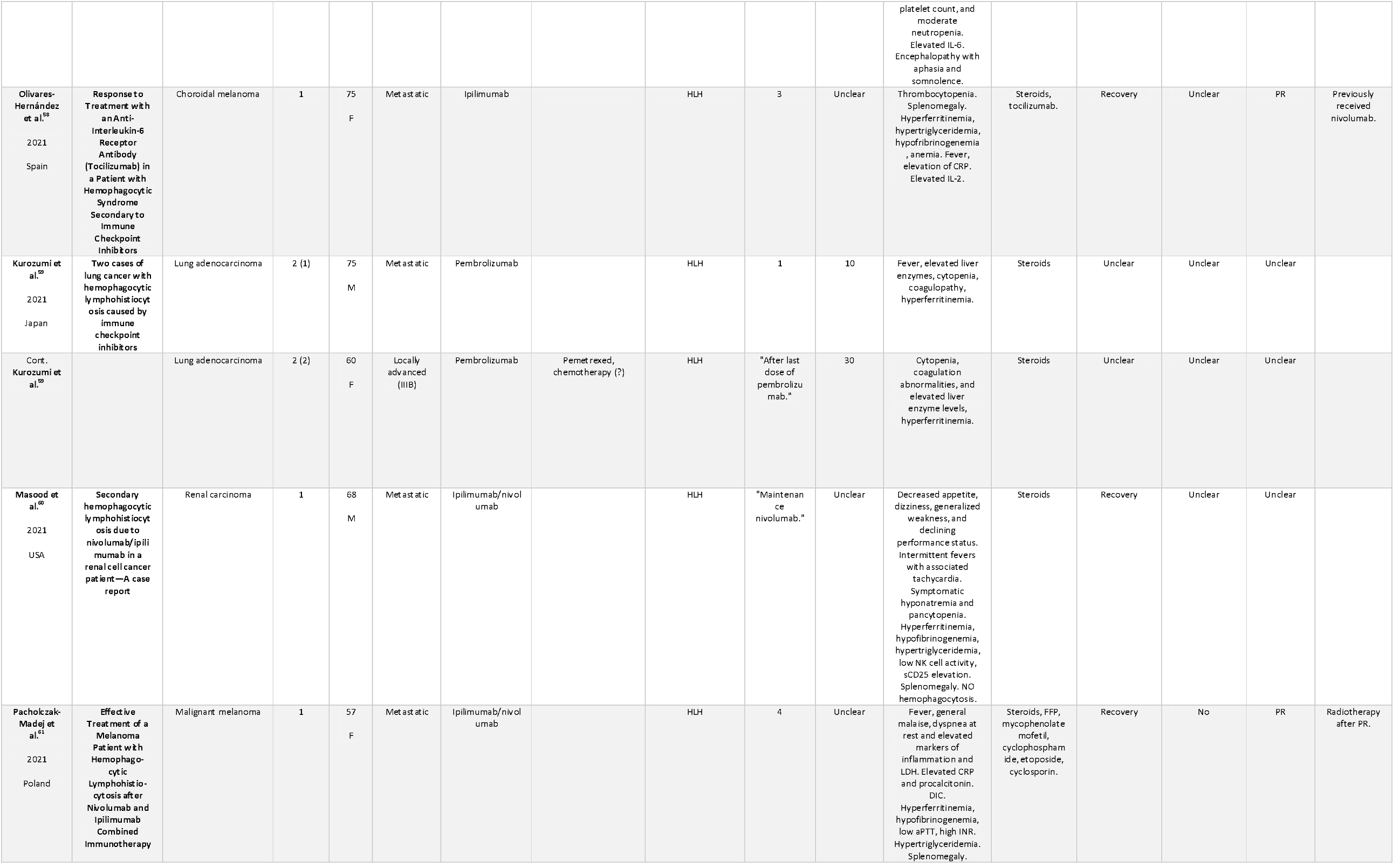
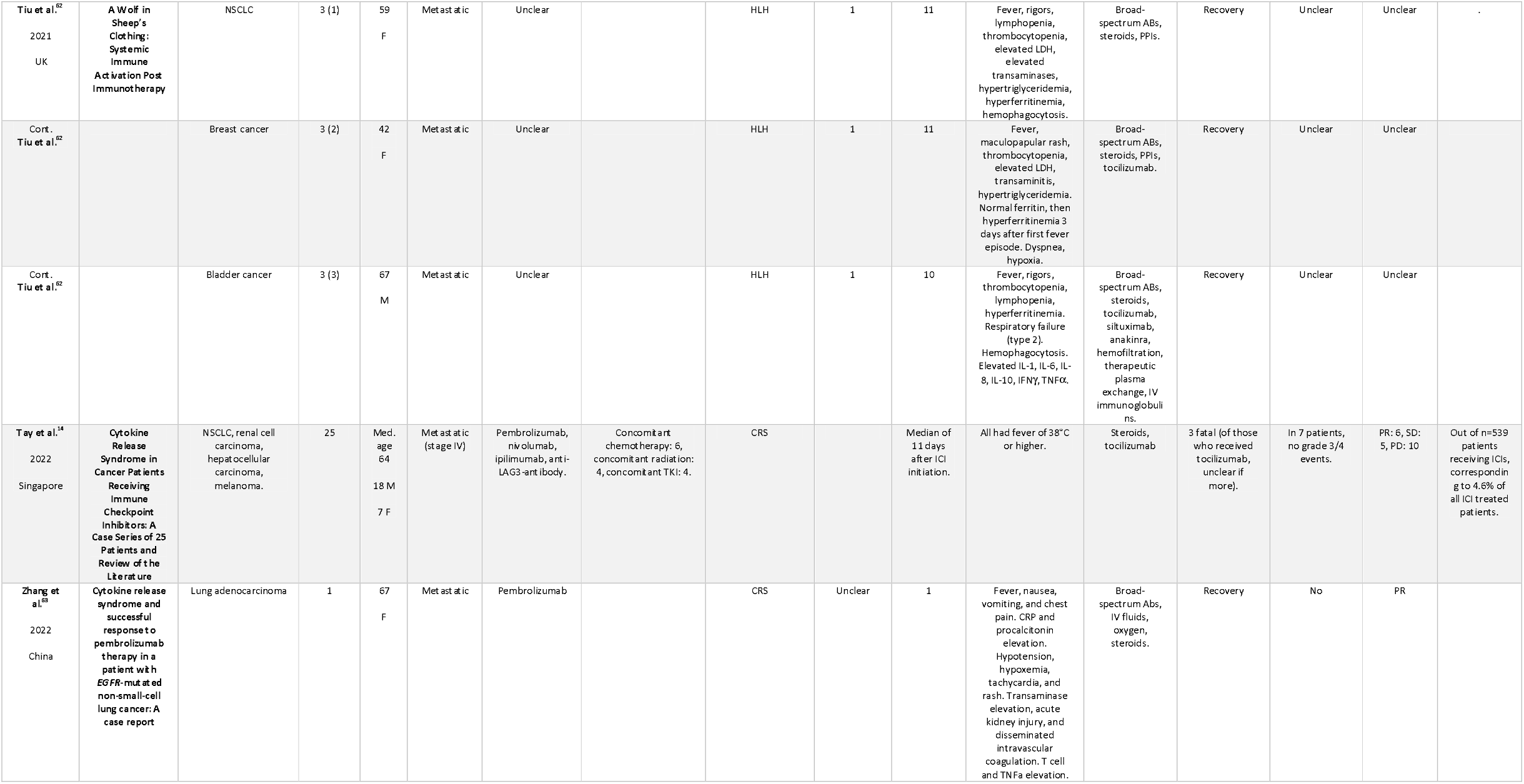
Studies retrieved in systematic review

The ICIs used included pembrolizumab (*n* = 21 studies) and nivolumab (*n* = 11 studies) as single agents, combined treatment with ipilimumab and nivolumab (n = 13), and, less frequently (*n* = 2 studies each), ipilimumab, sintilimab or atezolizumab monotherapy, and cemiplimab (*n* = 1). Often, ICIs were combined either with chemotherapeutical agents or other anticancer drugs, such as TKIs (i.e. cabozantinib or dabrafenib/trametinib), or drugs that were used prior to development of the hyperinflammatory syndrome(s).

In the individual reports, *n* = 15 patients were diagnosed with CRS, *n* = 34 with HLH, and *n* = 6 were described as having related hyperinflammatory conditions such as capillary leak syndrome with high fever, or systemic inflammatory response syndrome (SIRS). The most frequently reported underlying malignancies in individual case reports were malignant melanoma (*n* = 21 individual reports) and lung cancer (*n* = 14 individual reports). The average age of the patients in the individual reports was 59 years. In individual reports that reported the patients’ sex, *n* = 35 patients were males, and *n* = 20 patients were females. The highest frequencies of male patients were also reported in the largest case series from two centers in Singapore^14^ (*n* = 18 males, *n* = 7 females), and in a World Health Organization (WHO) pharmacovigilance database queried for HLH^15^ (*n* = 29 males, *n* = 9 females) and CRS, respectively^16^ (*n* = 34 males, *n* = 21 females). Most studies (n = 46 studies out of 49) comprised data on patients with metastatic disease (**Table 1**). Time from ICI infusion to onset of symptoms varied from hours to one month, with a median of *n* = 9 days. In total, *n* = 42 studies on n = 56 individual patients (75%) reported recovery from ICI-induced irAEs, while *n* = 5 cases (out of 56, corresponding to 8.9%) were fatal. Of those, *n* = 3 cases were reported to have HLH, and *n* = 2 had CRS, as per the authors’ assessment. Overall, *n* = 42 patients from individual reports were reported to have recovered from their hyperinflammatory complications. Reports from pharmacovigilance data were incomplete for outcome data; a study including *n* = 25 patients from Singapore reported *n* = 3 fatal cases (0.12 %).

Treatments for CRS/HLH varied significantly, and included different types of corticosteroids in almost all cases, and tocilizumab in *n* = 14 cases. Various other drugs were used in some patients, including etoposide in combination with dexamethasone, an established HLH treatment^17,18^, as well as intravenous immunoglobulins, plasmapheresis, mycophenolate mofetil, and tacrolimus. Often, these were combined, at least initially, with antibiotic therapy because of suspected sepsis (**Table 1**).

## Discussion

CRS and HLH are potentially life-threatening side effects of treatment with ICIs; however, they are relatively rare and therefore are challenging to study. In this systematic review, we provide a comprehensive picture of the reported cases in the literature to date. Our results underscore the notion that hyperinflammatory syndromes are rare, often treatable, and rarely fatal (**Table 1**).

It is interesting to note the predominance of male patients in both case reports and small case series, as well as in pharmacovigilance databases. This might partly be driven by a higher proportion of men among lung cancer patients^64^, which represent the second largest tumor group in the identified studies, and partly by sex differences in the immune response^65^. Interestingly, pharmacovigilance data suggest that the outcome of CRS is more favorable in females than in males^16^, warranting further research into sex differences in response to treatment and in side effect profiles of ICIs.

Wider indications and lower costs have continuously led to higher numbers of patients being treated with ICIs. Hence, the clinical need to understand rare side effects has increased, and higher patient numbers allow more informed clinical decisions. A common clinical dilemma in patients receiving ICIs is whether to continue treatment despite severe, and potentially life-threatening irAEs. Key factors in this decision-making process are the risk of recurrence of a given irAE, the anticipated severity in case of recurrence, as well as its treatability. A recent study has suggested that IL-6 blockade given in parallel with ICIs ameliorates irAEs, while enhancing the antitumoral effect of ICIs^66^. Earlier experimental data had already hinted at the potential benefit of adding IL-6 inhibitors to ICIs in mouse models of pancreatic cancer^67^ (which is largely resistant to ICI therapy), as well as hepatocellular carcinoma^68^. Together, these data suggest that IL-6 blockade does not abrogate, but rather enhances, the activation of a beneficial antitumoral immune response, providing a potential oncological rationale for combining IL-6 inhibition and ICIs. Results from CAR-T cell therapy in hematological malignancies support the lack of antagonistic effects of IL-6 antagonists in combination with ICIs; for example, in refractory large B-cell lymphoma, response rates to CAR T cell therapy were independent of the use of concomitant tocilizumab to treat CRS^69^.

Our review of the literature revealed that IL-6 inhibitors have successfully been used in cases of hyperinflammatory irAEs upon ICI treatment (**Table 1**). Given the evidence that combined anti-IL-6/ICI treatment enhances the antitumoral effect and is highly effective for CRS, an important open question is whether a rechallenge after mild CRS/HLH that responded well to anti-IL-6 treatment or corticosteroids should be considered. As shown in **Table 1**, only a few case reports exist on ICI rechallenge after CRS or HLH: One patient with metastatic oropharyngeal squamous cell carcinoma developed HLH after cycle 14 of pembrolizumab and was successfully treated with dexamethasone and etoposide^49^. Eight months after this episode, during which no treatment was given, tumor progression was noted, and the patient was restarted on pembrolizumab without signs of recurrent HLH after five additional cycles^49^. Another patient with a locally advanced lung adenocarcinoma developed CRS after the first cycle of pembrolizumab as first-line treatment, and was successfully treated with methylprednisolone^31^. Pembrolizumab was restarted while the patient was on low dose oral corticosteroids; the patient experienced no relapse of CRS and showed a near-complete tumor response to pembrolizumab^31^.

As a higher tumor burden has been shown to correlate with increased inflammation parameters and cytokine levels^70^, it can be hypothesized that pre-treatment tumor burden is associated with the risk of developing irAEs. Indeed, studies of CAR T cell treatment for B cell acute lymphoblastic leukemia (B-ALL) have shown a significant correlation between severe CRS and a high tumor burden^71,72(p19)^. In the largest case series published containing *n* = 25 cases, *n* = 7 were cases after rechallenge with ICIs; none of these had grade 3 or 4 CRS^14^, which lends some support for continuous ICI treatment despite CRS and could be appropriate in selected cases, as the risk of aggravated side effects when re-exposing patients to ICIs could be low.

While we mostly found case reports, two studies reported pharmacovigilance data from the global WHO database, and two reports queried the Registry of Severe Adverse Reactions to Immunomodulatory Antibodies used in Oncology (REISAMIC), a database for ICI-related irAEs in France^24,45^. The first report on REISAMIC data included *n* = 16 patients with “fever reaction” to ICI treatment, and based on their analysis of these cases, the authors concluded that this irAE can “usually be controlled with a short course of corticosteroids”. The second report also queried two other French databases specifically for HLH cases, and identified *n* = 5 patients with HLH, one of which was fatal. In *n* = 3 of the five cases, rechallenge of ICI was reported, in *n* = 2 cases without recurrence of fever of HLH^45^. One of the HLH cases was identified in REISAMIC among *n* = 745 patients included “at a single center between 2014 and 2019”, suggesting that HLH indeed is a rare event. The rarity of HLH is also supported by the analysis of WHO pharmacovigilance data: 49,883 ICI-related adverse events were retrieved from the WHO database VigiBase on a search conducted in September 2018, *n* =38 of whom corresponded to HLH, and *n* = 34 were directly linked to ICI treatment, usually developing more than six weeks after ICI treatment. Interestingly, the rate of other irAEs was below 20%^15^. The same group of authors queried VigiBase for CRS as of January 2020. They found *n* = 58 reports likely corresponding to CRS among a total of 80,700 reports on ICI-related adverse events, of which *n* = 43 were definitely related to ICIs, and which occurred a median of approximately four weeks after initiation of ICI treatment^16^. Two of those cases were fatal. Finally, a recent study presented a case series collected at two hospitals in Singapore between February 2014 and January 2021. They found that *n* = 25 out of a total of *n* = 539 patients that had received ICI developed CRS, which is a considerably higher frequency than suggested by the pharmacovigilance data, and the reason for this difference is unclear. In this cohort, *n* = 7 patients with low grade CRS were rechallenged with ICIs and did not relapse. A total of *n* = 3 cases had fatal CRS despite tocilizumab treatment^14^. The authors also suggested that time-to-fever-onset, low platelet count and high urea levels at CRS presentation might serve as indicators of a severe course ^14^.

Although several studies have reported an association between irAEs and improved treatment outcome^73,74^, it is still debatable whether ICI rechallenge after irAEs might be oncologically favorable. In accordance with current guidelines, the majority of patients with grade 3 or 4 irAEs will discontinue treatment with ICI permanently. The rarity of cases, resulting in cohorts of limited size, poses a challenge when addressing the question of ICI rechallenge. While recent studies have shown that rechallenge with ICI after irAE does not significantly improve overall survival^75^, others have concluded that irAEs upon rechallenge display milder toxicities, and suggested that rechallenge might be safe for most patients, depending on the type of irAE^76,77^. Furthermore, a meta-analysis of patients with non-small cell lung cancer rechallenged with ICIs suggests that certain patients with disease progression during ICI discontinuation might benefit from ICI rechallenge^78^. Whether these data are applicable to ICI-related CRS remains to be seen.

In summary, the published data suggest that CRS and HLH are infrequent, potentially severe, but frequently treatable side effects of ICIs, and that rechallenge could be considered in selected cases, with IL-6 inhibition as an attractive preventive and therapeutic option.

## Data Availability

Data are available from the authors upon request.

## Declarations

### Availability of data and material

Data are available from the authors upon request.

### Authors’ contributions

LLL and MG conceived the study and performed the literature search together with the librarians of Karolinska Institute’s library. LLL, MS, and MG decided on study inclusion and collected the data for the systematic review. LLL, MG, and UH collected and interpreted the data from the clinical case. MG wrote the manuscript, LLL, MS, and UH edited the manuscript.

## Acknowledgements

The authors are grateful to Narcisa Hannerz and Sabina Gillsund of the Karolinska Institute’s Library, for expert help in conducting the systematic literature search. We are grateful to Marie Meeths for advice on the manuscript.

