## Supplementary Figure 1 for "Systemic inflammatory syndromes as life-threatening side effects of immune checkpoint inhibitors: Systematic review of the literature"

### 1. Medline

| Interface: Ovid MEDLINE(R) and Epub Ahead of Print, In-Process & Other Non-Indexed Citations and Daily<br><br>Date of Search: 26th October 2021<br><br>Number of hits: 444<br><br>Comment: In Ovid, two or more words are automatically searched as phrases; i.e. no quotation marks are needed | Field labels<br><br>• exp/ = exploded MeSH term<br>• / = non exploded MeSH term<br>• .ti,ab,kf. = title, abstract and author keywords<br>• adjx = within x words, regardless of order<br>• * = truncation of word for alternate endings |
|---|---|

Database(s): **Ovid MEDLINE(R) and Epub Ahead of Print, In-Process, In-Data-Review & Other Non-Indexed Citations and Daily** 1946 to October 25, 2021
Search Strategy:

| # | Searches | Results |
|---|---|---|
| 1 | Cytokine Release Syndrome/ | 1316 |
| 2 | Interleukin-1/ | 35705 |
| 3 | Interleukin-6/ | 67997 |
| 4 | Systemic Inflammatory Response Syndrome/ | 6384 |
| 5 | (cytokine* adj3 (storm* or syndrome*)).ti,ab,kf. | 6121 |
| 6 | ((inflammatory response or multi-system inflammatory or sepsis or septic or systemic inflammat*) adj3 syndrome*).ti,ab,kf. | 9329 |
| 7 | (hypercytokinemia* or hypercytokinaemia* or interleukin 1 or interleukin1 or interleukin i or il-1 or il1 or interferon beta 2 or interferon beta2 or interleukin b or interleukin hp1 or interleukin 6 or interleukin6 or IL6 or IL 6).ti,ab,kf. | 199037 |
| 8 | ((b-cell differentiation or b-cell stimulatory or b lymphocyte stimulating or hepatocyte stimulating or hybridoma or plasmacytoma) adj2 factor*).ti,ab,kf. | 710 |
| 9 | or/1-8 | 231624 |
| 10 | Macrophage Activation Syndrome/ | 557 |
| 11 | Lymphohistiocytosis, Hemophagocytic/ | 3250 |
| 12 | (macrophage activati* adj2 syndrome*).ti,ab,kf. | 1234 |
| 13 | ((Erythrophagocyt* or hemophagocyt*) adj2 (histiocyt* or hymphohistiocyt* or lymphohistiocyt* or reticulosis or syndrome*)).ti,ab,kf. | 5341 |
| 14 | or/10-13 | 7028 |

Karolinska Institutet

| | | |
|---|---|---|
| 15 | 9 or 14 | 237794 |
| 16 | Immune Checkpoint Inhibitors/ | 2924 |
| 17 | Ipilimumab/ | 2300 |
| 18 | Nivolumab/ | 3587 |
| 19 | (atezolizumab or avelumab or cemiplimab or durvalumab or pembrolizumab).nm. | 3755 |
| 20 | (atezolizumab or avelumab or cemiplimab or durvalumab or ipilimumab or nivolumab or pembrolizumab).ti,ab,kf. | 13171 |
| 21 | ((CTLA-4 or CTLA4 or Cytotoxic T-Lymphocyte-Associated Protein 4 or checkpoint or PD-1 or PD1 or PD-L1 or PDL1 or Programmed Death-Ligand 1 or Programmed Cell Death Protein 1) adj3 (block* or inhibit*)).ti,ab,kf. | 30253 |
| 22 | (Anti-CTLA-4 MAb or strentarga or yervoy).ti,ab,kf. | 169 |
| 23 | (anti-PDL1 or anti-PD L1 or monoclonal antibody mpdl 3280a or monoclonal antibody mpdl3280a or tecentriq).ti,ab,kf. | 2132 |
| 24 | (bavencio or imfinzi or keytruda or lambrolizumab or libtayo).ti,ab,kf. | 135 |
| 25 | or/16-24 | 38419 |
| 26 | 15 and 25 | 527 |
| 27 | 26 not (animals not humans).sh. | 465 |
| 28 | limit 27 to yr="2013 -Current" | 444 |

#### 2. Embase

| Interface: embase.com<br><br>Date of Search: 26th October 2021<br><br>Number of hits: 1,579<br><br>Comment: Emtree is the controlled vocabulary in Embase | Field labels<br><br>• /exp = exploded Emtree term<br>• /de = non exploded Emtree term<br>• ti,ab,kw = title, abstract and author keywords<br>• NEAR/x = within x words, regardless of order<br>• * = truncation of word for alternate endings |
|---|---|

| No. | Query | Results |
|---|---|---|
| #1 | 'cytokine storm'/exp | 10555 |
| #2 | 'interleukin 1'/de | 67261 |
| #3 | 'interleukin 6'/de | 279870 |
| #4 | 'systemic inflammatory response syndrome'/de | 13389 |
| #5 | (cytokine* NEAR/3 (storm* OR syndrome*)):ti,ab,kw | 9621 |
| #6 | (('inflammatory response' OR 'multi-system inflammatory' OR sepsis OR septic OR 'systemic inflammat*') NEAR/3 syndrome*):ti,ab,kw | 14236 |
| #7 | hypercytokinemia*:ti,ab,kw OR hypercytokinaemia*:ti,ab,kw OR 'interleukin 1':ti,ab,kw OR interleukin1:ti,ab,kw OR 'interleukin i':ti,ab,kw OR 'il-1':ti,ab,kw OR il1:ti,ab,kw OR 'interferon beta 2':ti,ab,kw OR 'interferon beta2':ti,ab,kw OR 'interleukin b':ti,ab,kw OR 'interleukin hp1':ti,ab,kw OR 'interleukin 6':ti,ab,kw OR interleukin6:ti,ab,kw OR il6:ti,ab,kw OR 'il 6':ti,ab,kw | 274664 |
| #8 | (('b-cell differentiation' OR 'b-cell stimulatory' OR 'b lymphocyte stimulating' OR 'hepatocyte stimulating' OR hybridoma OR plasmacytoma) NEAR/2 factor*):ti,ab,kw | 787 |
| #9 | #1 OR #2 OR #3 OR #4 OR #5 OR #6 OR #7 OR #8 | 401096 |
| #10 | 'hemophagocytic syndrome'/exp | 11879 |
| #11 | ('macrophage activati*' NEAR/2 syndrome*):ti,ab,kw | 2383 |
| #12 | ((erythrophagocyt* OR hemophagocyt*) NEAR/2 (histiocyt* OR hymphohistiocyt* OR lymphohistiocyt* OR reticulosis OR syndrome*)):ti,ab,kw | 8173 |
| #13 | #10 OR #11 OR #12 | 13379 |
| #14 | #9 OR #13 | 412207 |
| #15 | 'immune checkpoint inhibitor'/de | 8085 |
| #16 | 'avelumab'/de | 3790 |
| #17 | 'atezolizumab'/de | 8825 |
| #18 | 'cemiplimab'/de | 724 |
| #19 | 'durvalumab'/de | 6008 |

Karolinska Institutet

| | | |
|---|---|---|
| #20 | 'ipilimumab'/de | 17977 |
| #21 | 'nivolumab'/de | 25266 |
| #22 | 'pembrolizumab'/de | 23295 |
| #23 | atezolizumab:ti,ab,kw OR avelumab:ti,ab,kw OR cemiplimab:ti,ab,kw OR durvalumab:ti,ab,kw OR ipilimumab:ti,ab,kw OR nivolumab:ti,ab,kw OR pembrolizumab:ti,ab,kw | 30924 |
| #24 | (('ctla-4' OR ctla4 OR 'cytotoxic t-lymphocyte-associated protein 4' OR checkpoint OR 'pd-1' OR pd1 OR 'pd-l1' OR pdl1 OR 'programmed death-ligand 1' OR 'programmed cell death protein 1') NEAR/3 (block* OR inhibit*)):ti,ab,kw | 55830 |
| #25 | 'anti-ctla-4 mab':ti,ab,kw OR strentarga:ti,ab,kw OR yervoy:ti,ab,kw | 316 |
| #26 | 'anti-pdl1':ti,ab,kw OR 'anti-pd l1':ti,ab,kw OR 'monoclonal antibody mpdl 3280a':ti,ab,kw OR 'monoclonal antibody mpdl3280a':ti,ab,kw OR tecentriq:ti,ab,kw | 5479 |
| #27 | bavencio:ti,ab,kw OR imfinzi:ti,ab,kw OR keytruda:ti,ab,kw OR lambrolizumab:ti,ab,kw OR libtayo:ti,ab,kw | 319 |
| #28 | #15 OR #16 OR #17 OR #18 OR #19 OR #20 OR #21 OR #22 OR #23 OR #24 OR #25 OR #26 OR #27 | 85919 |
| #29 | #14 AND #28 | 2602 |
| #30 | #29 NOT ([animals]/lim NOT [humans]/lim) | 2277 |
| #31 | #30 AND ('Conference Abstract'/it OR 'Conference Paper'/it) | 631 |
| #32 | #30 NOT #31 | 1646 |
| #33 | #32 AND (2013:py OR 2014:py OR 2015:py OR 2016:py OR 2017:py OR 2018:py OR 2019:py OR 2020:py OR 2021:py OR 2022:py) | 1579 |

Karolinska Institutet

### 3. Web of Science Core Collection

| | |
|---|---|
| Interface: Clarivate Analytics<br><br>Date of Search: 26th October 2021<br><br>Number of hits: 632 | Field labels<br><br>• TS/Topic = title, abstract, author keywords and Keywords Plus<br>• NEAR/x = within x words, regardless of order<br>• * = truncation of word for alternate endings<br><br>Note: sometimes “quotation marks” are needed for single search terms to avoid automatic term mapping (lemmatization). |

| | | |
|---|---|---|
| #1 | TS=(cytokine* NEAR/2 (storm* or syndrome*) ) | 6,944 |
| #2 | TS=(("inflammatory response" OR "multi-system inflammatory" OR "sepsis" OR "septic" OR "systemic inflammat*") NEAR/2 syndrome*) | 10,737 |
| #3 | TS=(hypercytokinemia* OR hypercytokinaemia* OR "interleukin 1" OR "interleukin1" OR "interleukin i" OR "il-1" OR "il1" OR "interferon beta 2" OR "interferon beta2" OR "interleukin b" OR "interleukin hp1" OR "interleukin 6" OR "interleukin6" OR "IL6" OR "IL 6") | 278,354 |
| #4 | TS=(("b-cell differentiation" OR "b-cell stimulatory" OR "b lymphocyte stimulating" OR "hepatocyte stimulating" OR "hybridoma" OR "plasmacytoma") NEAR/1 factor*) | 875 |
| #5 | TS=("macrophage activati*" NEAR/1 syndrome*) | 2,067 |
| #6 | TS=((erythrophagocyt* OR hemophagocyt*) NEAR/1 (histiocyt* OR hymphohistiocyt* OR lymphohistiocyt* OR "reticulosis" OR syndrome*) ) | 7,347 |
| #7 | #6 OR #5 OR #4 OR #3 OR #2 OR #1 | 300,364 |
| #8 | TS=(atezolizumab OR avelumab OR cemiplimab OR durvalumab OR ipilimumab OR nivolumab OR pembrolizumab) | 28,861 |
| #9 | TS=(("CTLA-4" OR "CTLA4" OR "cytotoxic t-lymphocyte-associated protein 4" OR "checkpoint" OR "PD-1" OR "PD1" OR "PD-L1" OR "PDL1 "OR "programmed death-ligand 1" OR "programmed cell death protein 1") NEAR/2 (block* OR inhibit*) ) | 37,463 |
| #10 | TS=("Anti-CTLA-4 MAb" OR "strentarga" OR "yervoy") | 159 |
| #11 | TS=("anti-PDL1" OR "anti-PD L1" OR "monoclonal antibody mpdl 3280a" OR "monoclonal antibody mpdl3280a" OR "tecentriq") | 3,108 |
| #12 | TS=("bavencio" OR "imfinzi" OR "keytruda" OR "lambrolizumab" OR "libtayo") | 150 |
| #13 | #12 OR #11 OR #10 OR #9 OR #8 | 55,381 |
| #14 | Refined by: PUBLICATION YEARS: ( 2021 OR 2015 OR 2020 OR 2014 OR 2019 OR 2013 OR 2018 OR 2017 OR 2016 ) Indexes=SCI-EXPANDED, SSCI, A&HCI, ESCI Timespan=1945-2021 | 670 |

Karolinska Institutet

| | | |
|---|---|---|
| #15 | #13 AND #7<br>Refined by: PUBLICATION YEARS: ( 2021 OR 2015 OR 2020 OR 2014 OR 2019 OR 2013 OR 2018 OR 2017 OR 2016 )<br>*Indexes=SCI-EXPANDED, SSCI, A&HCI, ESCI Timespan=1945-2021* | 632 |
